# Evolving Transmission Network Dynamics of COVID-19 Cluster Infections in South Korea: a descriptive study

**DOI:** 10.1101/2020.05.07.20091769

**Authors:** Yejin Kim, Xiaoqian Jiang

## Abstract

**Background:** Extensive contact tracing and testing in South Korea allows us to investigate the transmission dynamics of the COVID-19 into diverse local communities.

**Objective:** Understand the critical aspects of transmission dynamics in a different age, sex, and clusters with various activities.

**Methods:** We conducted a retrospective observational study with 3,127 confirmed cases’ contact tracing data from the Center for Disease and Prevention (CDC) of South Korea. We investigated network property concerning infected persons’ demographics and different infection clusters.

**Findings:** Overall, women had higher centrality scores than men after week four, when the confirmed cases rapidly increased. Older adults have higher centrality than young/middle-aged adults after week 9. In the infection clusters, young/middle-aged adults’ infection clusters (such as religious gatherings and gym facilities) have higher average path lengths and diameter than older adult’s nursing home infection clusters.

**Interpretation:** Some women had higher reproduction numbers and bridged successive transmission than men when the confirmed cases rapidly increased. Similarly, some older adults (who were not residents of nursing homes) had higher reproduction numbers and bridged successive transmission than young/middle-aged adults after the peak has passed. The young/middle-aged adults’ religious gatherings and group workout have caused long successive transmissions. In contrast, the older adults’ nursing homes were a small world where the transmissions within a few steps can reach out to many persons.

**Funding:** UT Startup award, UT STARs award, and Cancer Prevention Research in Texas, and National Institute of General Medical Sciences

**Research in context:** *Evidence before this study:* On May 1, 2020, PubMed query (“COVID-19” OR “SARS-nCoV-2” OR “novel coronavirus” OR “nCoV”) AND (“transmission network” OR “transmission dynamics” OR “transmission pattern” OR “centrality”) AND (“cluster” OR “community”) yield eight peer-reviewed papers. These papers have provided an evolving epidemiology and transmission dynamics via estimated reproduction number. However, most of them have focused on the entire system in one location and there was no comparison between transmission dynamics of different clusters.

*Added value of this study:* This study, to the best of our knowledge, is the first to compare the transmission dynamics of different cluster infections. We present the transmission dynamic with varying levels of granularity: entire country vs cluster infections as a longitudinal view. From the whole country-level analysis, we found that females have higher centrality (degree or betweenness) than males. From the cluster infection view, we found that young/middle-aged adults’ infection clusters (such as religious gatherings and gym facilities) have higher average path lengths and diameter than older adult’s nursing home infection clusters.

*Implications of all the available evidence:* This study sheds light on different transmission dynamics concerning demographics (age and sex) and diverse behavior in cluster infections. These findings are essential for planning tailored policies to diverse communities. Our analysis code is publicly available to adapt to newly reported cases.

## INTRODUCTION

Amid the global pandemic on coronavirus disease 2019 (COVID-19), epidemiologists promote containment strategies to prevent community transmission.(1) Contact tracing of susceptible persons and extensive testing in South Korea demonstrated as an example of successful containment. (1) South Korea reported the first confirmed cases on January 20, 2020, and the instances spiked from February 20 to 29, 2020.(2,3) As of mid-March, 2020, such containment strategies helped flatten the curve of new confirmed cases.(2)

The contact tracing and testing allow us to investigate different transmission dynamics into various local communities. Person-to-person transmissions can be represented as a transmission network, and this network facilitates investigation of topological dynamics in disease transmission and detection of super-spreaders.(4,5) Also, the transmission network can be divided into individual infection clusters that have different characteristics in terms of demographics and network topology. The network analysis has been widely studied in various infectious diseases such as Middle East Respiratory Syndrome (MERS),(4,5) Several Acute Respiratory Syndrome (SARS), (6)(6,7) tuberculosis,(8,9) and cholera.(10) Some COVID-19 studies have revealed evolving person-to-person transmission dynamics in China in a longitudinal view.(11,12) A genomic mutation study has investigated country-to-country transmission dynamics on a global scale.(13) A piece of critical missing knowledge is how person-to-person transmission differs across different demographics and also different super-spreading environments in cluster infections.

Thus, the objective of our study was to investigate network characteristics of person-to-person transmission in the different granularity: entire country and individual cluster infections. We provided interactive visualization in the code repository (https://github.com/yeiinjkim/covid19-transmission-network) for an intuitive understanding of the evolving transmission network. To the best of our knowledge, our work is first to compare transmission dynamics in cluster-wise.

## METHODS

### Data collection

We conducted a retrospective observational study with contact tracing data from the Center for Disease Controls and Prevention (CDC) of South Korea. The South Korea CDC actively performed testing, monitoring, and tracing contacts and disseminated the information to the public. We collected the contact tracing data prepared in the Data Science for COVID-19 Project in South Korea, (14) which enables researchers to access the data publicly. This data contains demographics, exposure type or travel history, confirmed/release date, underlying disease, deceased status. This study used 3,127 patient’s transmission data from January 20, 2020 to April 7, 2020, retrieved on April 8, 2020.

### Statistical Analysis

#### The evolving transmission network

We built a longitudinal transmission network using the transmission data. The transmission network consisted of nodes for the infected persons and directed edges from infectious nodes (infector) to infected nodes (infectee). The nodes and edges had timestamps at which the infectee’s infections are confirmed. This longitudinal network evolved over time as accumulating all previous nodes and edges.

#### Transmission network characteristics

We presented network characteristics in terms of centrality and path lengths:

- Centrality is a useful measure to investigate individual patients who shed the virus most (i.e., super-spreaders) over time. We analyzed network centrality with degree and betweenness. Node degree is the number of edges linked to the node in the network. Betweenness is the extent to which a node lies on the shortest paths between other nodes.(15) Nodes with high betweenness may have high control over transmission passing between nodes. We used betweenness to measure how much a patient contributed to the transmission as intermediate hosts. We defined a super-spreading person as a person with a degree > 3.5 for degree or top 5% betweenness values.(16) Note that the node degree was calculated based on all accumulated transmissions (whereas the reproduction number is generally calculated within the observation period).
- Path length can measure how far the transmission chain of successive cases can go. The path length is the number of nodes or steps in the shortest path from a source node to a target node. The average path length is the average value of the shortest path lengths between all pairs of nodes in the network. The longest path length (also known as diameter) is the maximum amount of the shortest path length.

#### Infection clusters

We divided the transmission network into subgraphs with connected nodes (i.e., the connected components). The connected components are an extended concept of infection cluster, as the connected components include all successive infected persons who are infected from the infection cluster. For briefly, we denoted the connected components as the infection cluster. During the contact tracing, it is expected that the index patient was unidentifiable in each infection cluster. To represent links among the nodes in the same cluster with an unknown index patient, we set a virtual index patient and we made edges from the virtual index patient to infected patients for each infection cluster.

## RESULTS

### The evolving transmission network

From January 20, 2020 to April 7, 2020, we identified a total 1,611 transmission from 3,127 patients (727 person to person; 884 persons in infection clusters with unknown index patient).

We presented demographic characteristics of patients and their transmission (Fig.1, Table 1) when newly confirmed cases rapidly increase (weeks 4, 5, 6) and after the recently confirmed cases decrease (after peak, week 10). Note that the mass cluster infection occurred at week 4 with an outbreak in religious gathering in Daegu.(17)

The demographic characteristics of patients and their transmission varied in different observation periods. Until week 4 (the confirmed positive cases were at a low level), male to female ratio was similar (53% male; 47% female). After week 4 South Korea reported more female cases than male ones. Also, female-to-female transmissions were more frequent than male-to-male transmissions (but there was no statistical power as chi-squared value of the contingency table=1.36, *p*-value=0.2430).

Throughout all observations, the largest age group was age 25-49. Although there was no reported transmission among similar aged children and adolescents (presumably due to strict school closure), this young age group sometimes infected their middle-aged parents. In week 6, South Korea reported an increased number of new confirmed cases from older adults, and the transmissions occurred mostly among the older adults.

**Table 1.** Demographics of newly confirmed cases.

|  | Feb 17 ~ Feb 24<br>(in week 4) |  | Feb 24 ~ March 2<br>(in week 5) |  | March 2 ~ March 9<br>(in week 6) |  | March 30 ~ April 6<br>(in week 10) |  |
| --- | --- | --- | --- | --- | --- | --- | --- | --- |
| Sex |  |  |  |  |  |  |  |  |
| Male | 90 | 53% | 123 | 43% | 74 | 32% | 71 | 45% |
| Female | 80 | 47% | 166 | 57% | 157 | 68% | 88 | 55% |
| Mean age (std) | 42.34 | 18.1 | 42.39 | 18.08 | 47.61 | 20.48 | 39.94 | 16.27 |
| Age group |  |  |  |  |  |  |  |  |
| 0-6 | 1 | 1% | 5 | 2% | 5 | 2% | 2 | 1% |
| 7-17 | 2 | 1% | 12 | 4% | 9 | 4% | 6 | 3% |
| 18-24 | 33 | 19% | 38 | 13% | 23 | 10% | 32 | 14% |
| 25-49 | 71 | 40% | 140 | 48% | 83 | 35% | 147 | 62% |
| 50-64 | 50 | 28% | 64 | 22% | 72 | 31% | 27 | 11% |
| >=65 | 20 | 11% | 35 | 12% | 44 | 19% | 23 | 10% |

**Figure 1.**
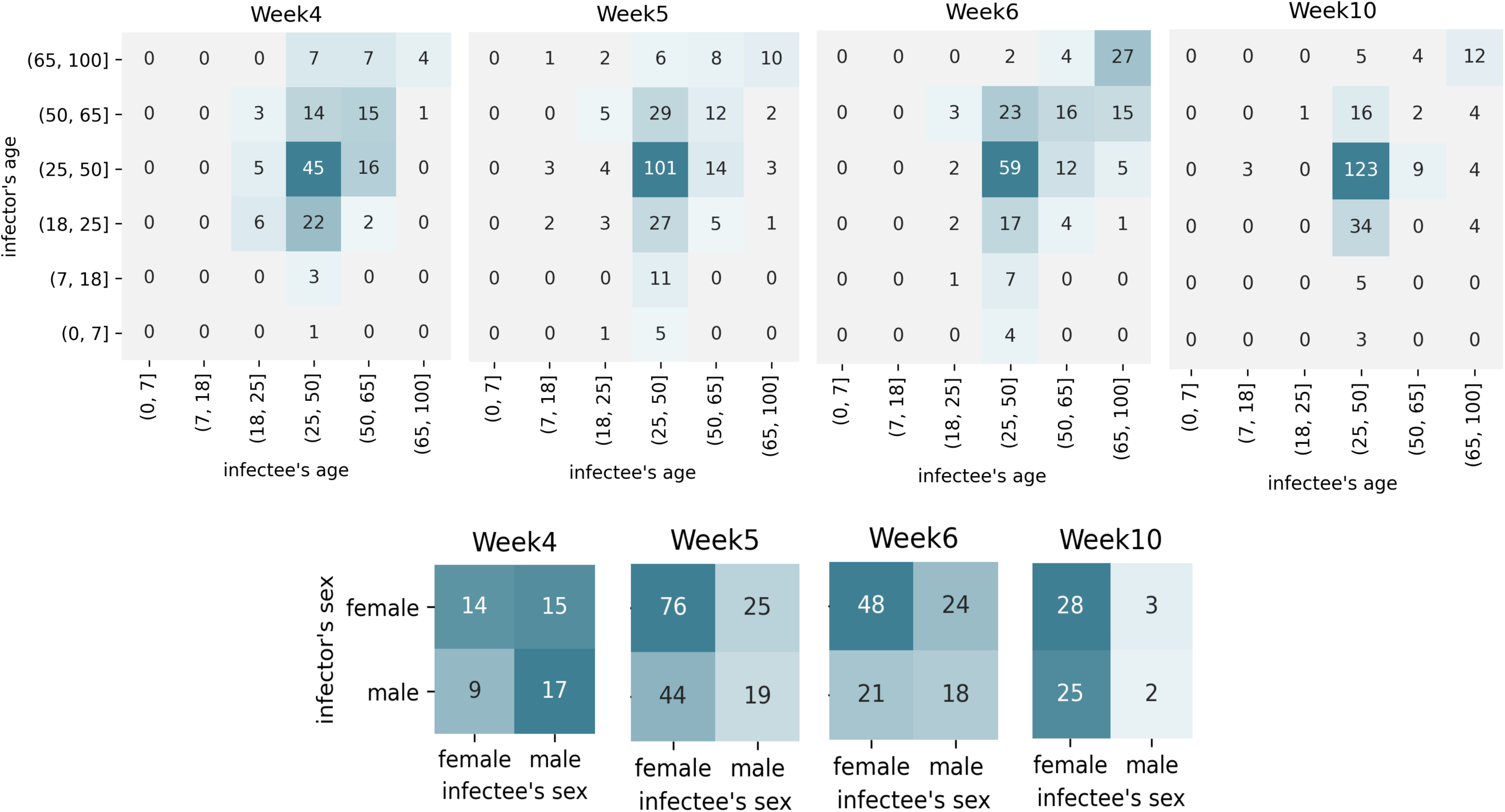
Demographics of newly confirmed cases and their infector.

We also presented the network property of the transmission network. The transmission network had a mean degree of 1.88 after aggregating all time windows. The system was getting sparse as new confirmed cases exponentially increased with unknown transmission paths (complete network visualization in Supplement 1).

**Figure 2.**
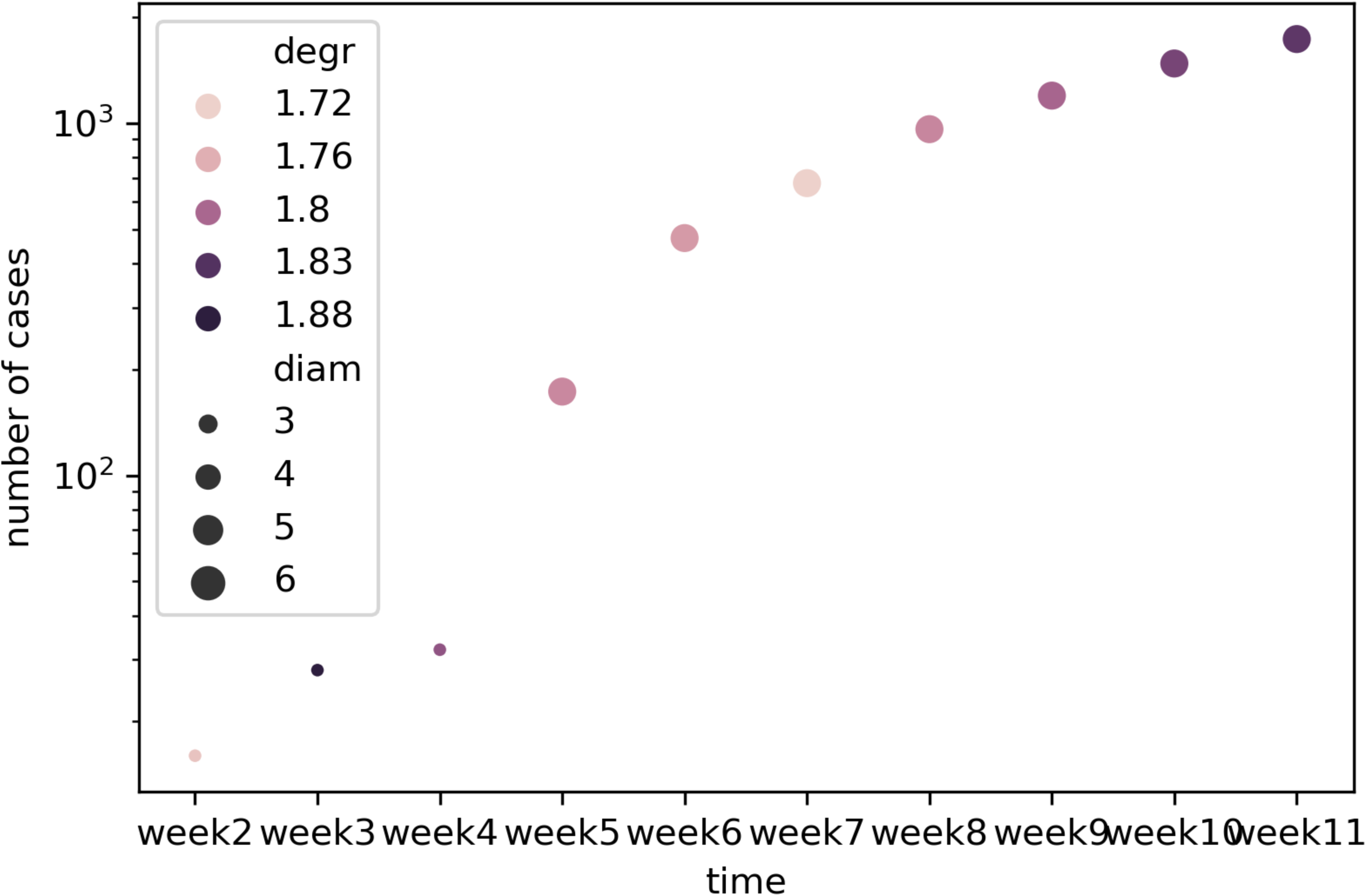
Transmission network characteristics of cumulative positive cases. Week 5 is when the local community spread starts with several clustered infections outbreak. degr=mean degree of nodes, diam=diameters (longest path length).

**Table 2.** Transmission network characteristics of cumulative positive cases until weeks 4, 5, 6, and 11. We focused on patients (nodes) that have edges (i.e., removed persons without known infection causes). Network characteristics were calculated from accumulated nodes and edge until the observation timestamp.

|  | Jan 20 ~ Feb 17<br>(until Week 4) | Jan 20 ~ Feb 24<br>(until week 5) | Jan 20 ~ March 2<br>(until week 6) | Jan 20 ~March 30<br>(until week 10) |
| --- | --- | --- | --- | --- |
| # Nodes | 32 | 173 | 472 | 1478 |
| # Edges | 29 | 154 | 418 | 1348 |
| Density | 0.0292 | 0.0052 | 0.0019 | 0.0006 |
| Mean degree | 1.8125 | 1.7804 | 1.7712 | 1.824 |
| Mean betweenness | 7.0564 *1e-4 | 2.2795 *1e-5 | 2.7276 * 1e-6 | 1.4728 *1e-7 |
| Diameter | 3 | 6 | 6 | 6 |
| Average path length | 1.7540 | 1.4633 | 1.4934 | 1.4728 |

### The centrality of individual patients

We further investigated the distribution of the network centrality concerning infected persons’ sex and race (animated plot in Supplement 2). We computed the statistical significance on the difference of centrality distribution using the Mann-Whitney-U test.(18)

As a result, we found that women have higher centrality scores than men as the confirmed cases exponentially increase around week 5 to 6 (Fig. 3). During the first four weeks after the first case, the number of instances slowly increased, and men tend to have higher degrees and betweenness. Women, however, started to have higher degrees and betweenness after week 5. We used the Mann-Whitney-U for hypothesis testing on whether super-spreading women have higher centrality values than super-spreading men. The *p*-values were statistically significant after week 6 for degree (*p*-value=0.0156) and week 11 for betweenness (*p*-value=0.0402).

We found that older adults have higher centrality than young/middle-aged adults after the peak has passed around week 9 (Fig. 3). After week 9, more older adults had higher degree and betweenness values than young/middle-aged adults. We used the Mann-Whitney-U for hypothesis testing on whether the super-spreading older adults have higher centrality than super-spreading young adults. The *p*-values were statistically significant after week 9 for degree (*p*-value=0.0413). Note that there was a time lag between degree and betweenness as the betweenness increases after successive transmission occurs.

**Figure 3.**
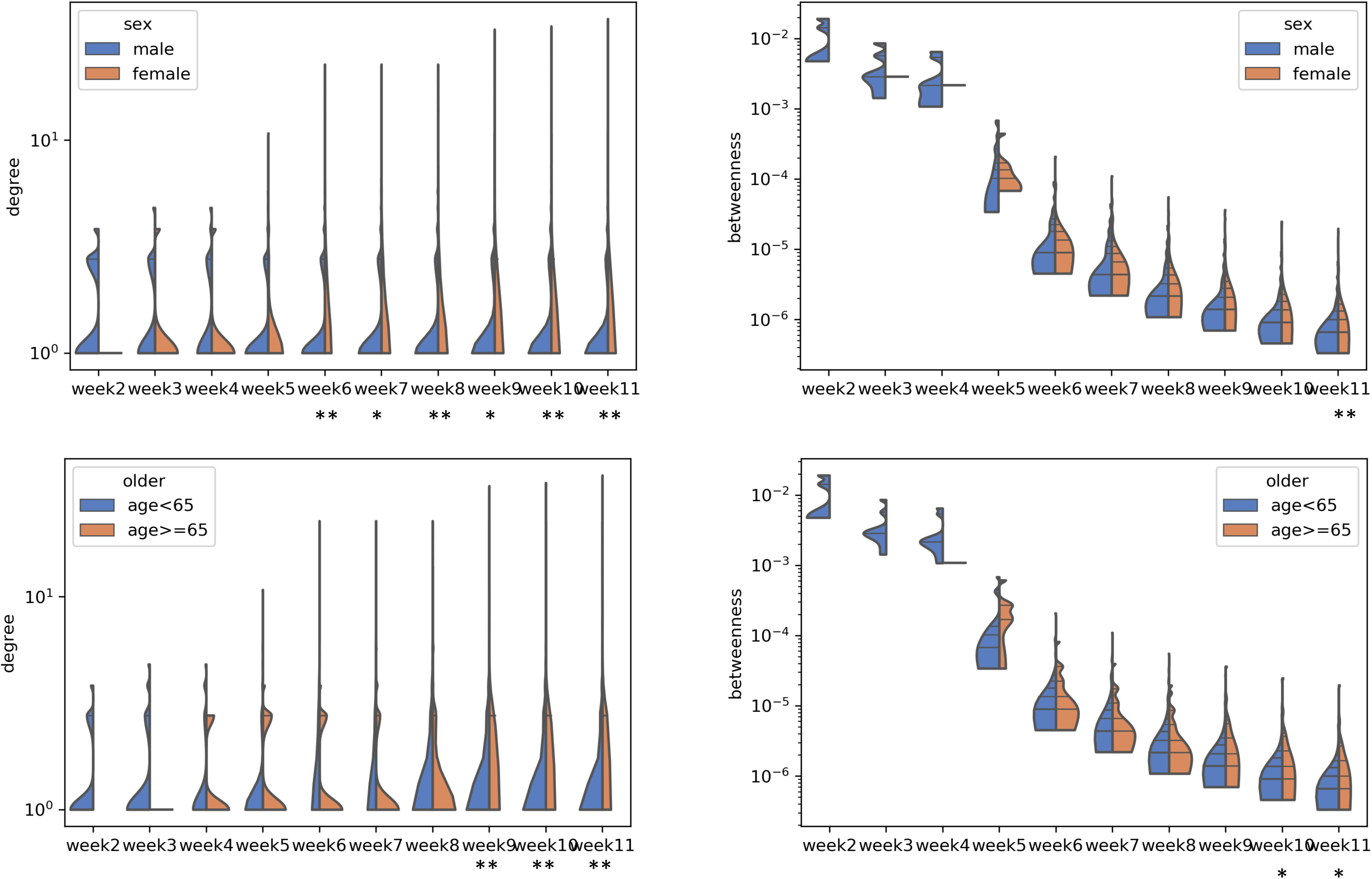
Centrality distribution concerning sex and age. Left: degree, Right: betweenness, Up: sex, Down: age>=65. * if *p*-value <0.1; ** if *p*-value<0.05 for the Mann-Whitney-U test. Note that overall betweenness score decreased as the number of confirmed patients exponentially increased with unknown transmission path and the transmission network became sparse.

### Infection clusters

There were a total of 147 clusters (defined as connected components) and we selected 12 clusters that have at least 20 patients involved. These clusters turned out to be related to religious gatherings, gym facilities, nursing homes, and customer call center office (Table 3, Fig. 4). The clusters’ network visualization is available at Supplement1. Overall, the top four largest clusters were related to *Guro* customer call center, *Shincheonji* Church, *River of Grace Community* Church, *and* gym facilities in *Cheonan.* A common characteristic of the four largest clusters was that they all had higher women rates (72%, 55%, 60%, and 72%, respectively). Clusters with the smallest path lengths were all related to nursing homes (*Gunpo Nursing Home, Cheongdo Daenam Hospital Psychiatric Ward, and Bongwha Nursing Home*). Clusters with the most considerable path length were related to Gym facilities *in Cheonan* and *Shincheonji* Church. We observed that similar activities share similar path lengths (three nursing homes had the path length of 1.91-1.94; four churches had the path length of 2.42-2.94).

**Table 3.** Cluster infection characteristics as of April 7, 2020. Elderly population % were general statistics of the city where the cluster infection occurs.

| Cluster | Network property |  |  |  | Demographics |  |  |  |
| --- | --- | --- | --- | --- | --- | --- | --- | --- |
|  | Cluster size | Path length | Avg. Degree | Diameter | Male % | Mean age (yrs) | Elderly population % in the province | Fatality % |
| Imported cases* | 575 | 2.47 | 2.00 | 6 | 59 | 35.76 | N/A | 0.00 |
| Religious gatherings |  |  |  |  |  |  |  |  |
| *Shincheonji* Church | 149 | 2.94 | 1.99 | 5 | 45 | 40.86 | 18.17 | 0.67 |
| *River of Grace* Community Church | 67 | 2.42 | 1.97 | 4 | 40 | 45.90 | 13.49 | 0.00 |
| *Onchun* Church | 43 | 2.68 | 1.95 | 3 | 49 | 34.50 | 18.01 | 0.00 |
| *Dongan* Church | 29 | 2.65 | 1.93 | 3 | 62 | 36.02 | 17.05 | 0.00 |
| Nursing homes |  |  |  |  |  |  |  |  |
| *Gunpo-si* Nursing Home | 22 | 1.91 | 1.91 | 1 | 18 | 77.55 | 12.68 | 0.00 |
| *Cheongdo Daenam Hospital* Psychiatric Ward | 22 | 1.91 | 1.91 | 1 | 73 | 50.62 | 35.72 | 31.82 |
| *Bonghwa Pureun* Nursing Home | 32 | 1.94 | 1.94 | 1 | 9 | 69.74 | 35.26 | 3.13 |
| Gym facilities |  |  |  |  |  |  |  |  |
| Gym Facility in *Cheonan-si* | 63 | 3.41 | 1.97 | 4 | 29 | 38.21 | 10.49 | 0.00 |
| *Cheonan/Asan-si* | 35 | 2.67 | 1.94 | 3 | 20 | 38.64 | 10.77 | 0.00 |
| Others |  |  |  |  |  |  |  |  |
| *Guro-gu* Customer Call Center | 164 | 2.80 | 1.99 | 4 | 28 | 44.67 | 14.30 | 0.00 |
| *Ministry of Oceans and Fisheries* | 30 | 2.12 | 1.93 | 2 | 73 | 40.33 | 9.59 | 0.00 |

\*Imported cases were represented as a single cluster to compare with other clusters.

**Figure 4.**
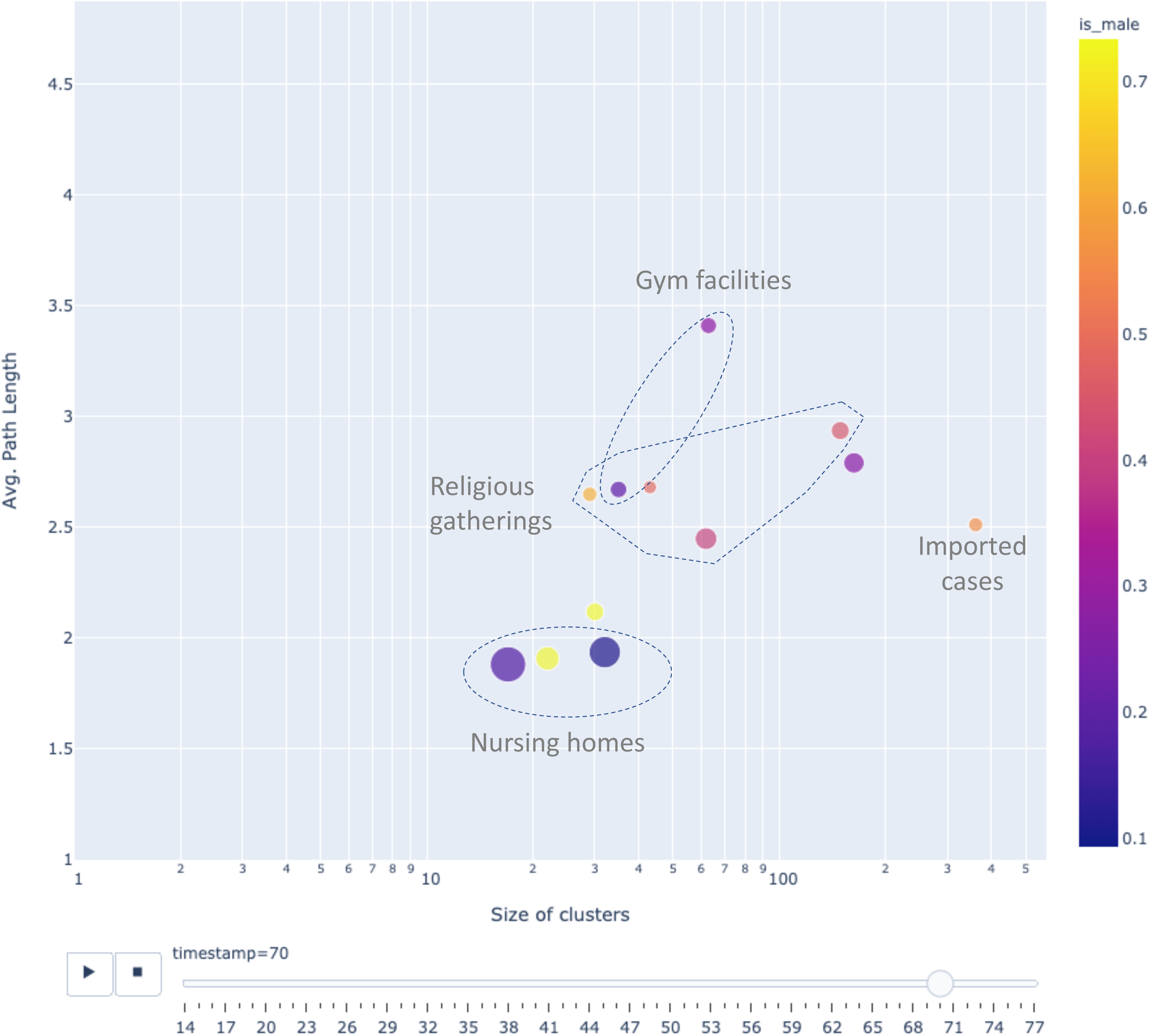
Evolving clusters over time. Avg. path length= averaged path lengths in one community; the size of clusters= number of infected patients in the clusters; Circle size = Mean age; Circle color=portion of men. Animated visualization is at Supplement 3.

## DISCUSSION

The objective of this study was to provide the evolving transmission dynamics of COVID-19 in South Korea. We investigated network property and demographic features from the whole transmission network and also clustered level.

The demographics of newly confirmed patients and their transmission provided an overview of how various demographic groups contribute to transmission. The main observation includes: i) In early days until week four most confirmed positives were imported cases who were male in their 40s; ii) women were more than 50% of newly confirmed cases after week 4; iii) there were more female-to-female transmissions than male-to-male transmissions; iv) children and adolescents infected their parents; and v) older adults infected other older adults. Further investigation is needed to obtain statistical power. We expect these observations can provide situational awareness to future contact tracing efforts.

The network characteristics of the entire system found the transmission network’s density and successive transmission lengths evolved over time. The transmission network was getting sparse as the number of newly confirmed cases rapidly increased. The sparsity limitation was inherited from the contact tracing containing the increasing number of unknown transmission paths in local communities. Thus our estimate on network property (degree, betweenness, diameter, and path length) should be interpreted as a lower bound.

The main finding from network centrality analysis was that women had higher centrality (in both degree and betweenness) than men had as the instances rapidly increased. This implies not only that women’s reproduction number was larger than men’s, but also that some women bridged the long successive transmissions. A possible explanation might be that men are more likely to die or critically ill than women, and consequently men have less opportunity to shed the virus.(19) Note that the higher portion of confirmed cases does not necessarily cause higher centrality.

The second finding is that older adults had high centrality after the peak has passed around week 9. Note that these older adults with high reproduction numbers were not related to nursing homes. Older adults are more vulnerable to critical illness.(20) Based on our findings, they were not active super-spreaders in the early days. Still, after the virus propagated into local communities, the older adults were more likely to be infected by the virus and be super-spreaders.

A breakdown into smaller infection clusters allowed us to find common network characteristics within similar activities. There were three major types of clusters: religious gatherings, gym facilities, and nursing homes. The religious gathering and gym facilities clusters had high path length and diameter. These clusters mainly consisted of young/middle-aged adults. This observation may imply that the active behavior (e.g., physically close gatherings in church, active group workout in closed facilities) of young/middle-aged adults can spread further away to other communities. Thus, restricting trips to other cities would be an adequate policy to prevent the spread effectively.

In contrast, the nursing home type clusters consist of older adults with low path lengths and low diameter. Each nursing home cluster grew fast with relatively short path length, implying that the clusters might have a small-world property that spread the virus rapidly as the majority of nodes were reachable in a small number of steps. Among the three nursing homes, *Cheongdo Daenam Hospital Psychiatric Ward* had the highest fatality, 31.82%. This closed psychiatric ward has many long-term patients with underlying diseases.(17)

Apart from the above local community infections, imported cases were also consistently increasing during all observation periods. The mean age of the imported patients was 42 years old at week two and consistently decreased to 35.6 years old until week 11. Men took more than 50% of the imported cases for all observation periods (from 71% to 59% at week two and week 11).

This study’s limitation mainly comes from the sample bias in contact tracing data. The contact tracing of the rapidly evolving infectious disease inevitably contains case ascertainment biases, non-homogenous sampling over time and location, and uncontrolled correlation.(11) A substantial number of cases were missing (officially confirmed cases from Korea CDC was more than 8,000 as of April). Each local province government performs and reports the contact tracing, but the difficulty of the instances or granularity of reporting varies from each local government. For example, the import cases were more likely to be tracked well, but the mass infection cluster in *ShinCheonji* Church has not been carefully investigated.

Despite the limitations, our descriptive study can provide an overview of evolving transmission dynamics in the country and infection clusters for the rapidly emerging infectious disease. Our findings suggest a different level of reproduction and successive transmission chain over sex, age, and cluster activities. Our analysis code is publicly available to adapt to newly reported cases.

## Data Availability

Data and analysis results are publicly available

## Acknowledgments

We collected the contact tracing data prepared in the *Data Science for COVID-19 Project* in South Korea. Y.K. and X.J. were partly supported by the UT Startup award, UT STARs award, and Cancer Prevention Research in Texas (CPRIT RR180012), and National Institute of Health (NIGMS R01GM124111).

## Supplementary Materials

Supplement 1: Network visualization for cluster infections

Supplement 2: Animated degree distribution concerning age and sex

Supplement 3: Animated evolving transmission dynamics of clusters

## Author Contributions

YK collected data;

YK and XJ performed statistical analysis; and

YK and XJ prepared the manuscript.

